# *“You’re too young to have an eye problem”*: Reasons for nonadherence to referrals for follow-up eye care for schoolchildren in Nigeria -- a descriptive qualitative study

**DOI:** 10.1101/2021.06.08.21258336

**Authors:** Lynne Lohfeld, Christine Graham, Anne Effiom Ebri, Nathan Congdon, Ving Fai Chan

**Affiliations:** Centre for Public Health, School of Medicine, Dentistry and Biological Sciences, Queen’s University Belfast, Northern Ireland, UK; Clinical and Epidemiological Eye Research Center, Wenzhou Medical University, Wenzhou, China; Nelson R. Mandela School of Medicine, KwaZulu Natal, Durban, South Africa; Brien Holden Vision Institute Foundation (Africa) Trust, Durban, South Africa; Zhongshan Ophthalmic Centre, Sun Yat-sen University, Guangzhou, China; Orbis International, New York, NY, USA; College of Health Sciences, University KwaZulu Natal, Durban, South Africa

**Keywords:** children, vision impairment, refractive error, spectacles, barriers, qualitative research, Nigeria

## Abstract

**Study objectives:** to identify reasons for non-adherence to referrals for follow-up eye care after children fail a school vision screening test.

**Methods:** Ten focus groups were held with parents or guardians (‘parents’) of children who had not adhered to the referral for further care in Cross River State, Nigeria, in 2019. Data from verbatim transcripts were analysed deductively using topics from the interview guide plus ‘Other’ to capture unanticipated results. Analysts followed procedures for Qualitative Content Analysis plus a modified Framework Method to identify overarching themes and barriers that are both highly salient (most frequently mentioned) and relevant (discussed in at least half of all groups).

**Results:** Three themes identified in the data are 1) modifiable barriers (key among them being parental beliefs and problems with the referral letter), 2) contextual factors (parents’ situation, attitudes towards children and beliefs about care) and 3) participants’ recommendations to improve the child eye care program (educate the general public and correct parents’ misconceptions). Many of the findings echoed those from previous studies conducted in both low-and-middle income countries (LMICs) and high-income countries (HICs).

**Conclusion:** This study went beyond identifying modifiable barriers to also identify contextual factors and what parents recommend be done to improve vision care for children in Cross River State, Nigeria. If acted on, these findings may increase acceptance and uptake of eye care services that can promote sustainability and spread of the program to other parts of Nigeria and/or Africa.

## Introduction

Approximately 17.5 million children have vision impairment and 1.4 million are blind globally, with most of them living in low- and middle-income countries (LMICs). [1-3] Uncorrected refractive errors (UREs) are the leading cause of childhood vision impairment (VI) in much of the world, including Africa, [4,5] with myopia the most common type of refractive error (RE). [6] Nigeria, where the present study was conducted, is home to nearly one-quarter of Africa’s visually impaired or blind children, [7] and has a RE prevalence rate of 5-8%. [8] Myopia and other forms of RE are asymptomatic, [9] making it more difficult to diagnose than other ocular problems with signs that children or their families can notice. This may be why in schoolchildren although cataract is the leading cause of blindness, RE accounts for double the number of blind person-years. [10]

VI can greatly affect people’s health and quality of life, [11] and is among the ten most frequent causes of disability in Africa. [12] UREs have a major economic impact on households and nations, [13] with an estimated annual global loss of US $269 billion due to URE-related VI as of 2009. [14] For these reasons, childhood blindness has been identified as a high-priority issue by the “World Health Organization’s VISION 2020: Right to Sight” program. [15]

As in many LMICs, Nigeria’s efforts to reduce UREs include school-based vision screening plus referral for further testing and care if children who fail the test, along with eye health promotion and access to free spectacles at designated child eye clinics. These services comprise the Comprehensive Child Eye Health in Nigeria program (CCEHiN), in effect from 2017 to 2020 in 11 of its 36 states. Currently in Cross River State, the Nigerian Ministry of Health is sponsoring a cross-subsidisation program whereby profits from selling mid-and high-price spectacles will be used to underwrite most of the cost of less expensive frames, making them affordable to low-income families.

The present study is part of a four-part investigation conducted in 2019 to provide evidence on what happens in Cross River State after children fail a school screening program, as described in another paper. [16] This paper reports on barriers to attending follow-up exams at a local hospital-based eye clinic for children who fail the school-based screening.

## Materials and methods

### Study design

This is a descriptive qualitative study [17,18] that aims to describe the views of parents or guardians (referred to as ‘parents’) on barriers to bringing schoolchildren who fail a vision screening test for further testing and care in local eye clinics.

### Sample and recruitment

The sample includes parents of children in four communities of Cross River State, Nigeria, where the subsidised eye care program is to be run. Eligible parents have a child who failed a school-based vision screening test (<6/12 in either eye) conducted by the CCEHiN program plus had not adhered to the referral for a follow-up eye exam. School health teachers identified potential participants and gave the list to the study research officer (RO). Two weeks before a scheduled focus group (FG), the RO visited potential participants in the area to describe the study, invite them to join a FG and leave them an information leaflet. About one week later the RO called to enrol participants, obtain informed oral consent and make arrangements for transportation to the interview venue. Consent was renewed in writing at the start of each FG. Half the interviews were conducted with parents affiliated with larger schools centrally located in each town (‘urban’), and the remainder were from smaller schools further away (‘rural’).

### Data collection

The interview guide was developed by VFC, AE and LL with input from staff in participating Cross River State eye clinics (see a copy of the guide in Supplementary File 1). Four fieldworkers, working in two teams (an interviewer and a note-taker), completed a three-day training program offered by VFC and AE in July 2019, which included pilot testing the interview guide with two groups of parents (pilot data not included in the analysed dataset). The fieldworkers had at least five years’ experience conducting community research; three of them were practising optometrists with O.D. (Doctor of Optometry) qualifications; and one note-taker had a college education. The two teams facilitated ten FGs from 15 October to 30 November 2019. Each session lasted 90-120 minutes and was audio-recorded with participants’ permission. The interviewers created verbatim written documents (transcripts) from the recordings, removing all identifying information and then compared audio and written versions to correct errors.

### Data analysis

Three team members (LL, VFC, CG) analysed the data deductively using the interview guide topics as a pre-set framework, plus inductively, added the heading ‘Other’ to capture unanticipated findings. The team followed procedures for Qualitative Content Analysis [19] and a modified version of the Framework Method [20], as follows. Working individually, the analysts read all the transcripts several times to familiarize themselves with the data and identify relevant statements (quotes). Next, they labelled each quote with an anonymous identifier (e.g., ‘FG1-M3’ = 3^rd^ male participant in FG #1). Then one of the team members created a table for each topic and populated it with relevant quotes from the first transcript. Starting with the first FG transcript, the analysts added relevant quotes to the tables, wrote brief summaries of the contents of each quote and used this to develop the initial codes + subcodes. Working in pairs, the analysts reviewed their codes until reaching consensus and then updated a codebook with the name, definition and example of each code/subcode. The analysts repeated this process until all data had been coded and the codebook was updated. A review of the data revealed no difference between urban- and rural-based parents’ responses so data from all ten FGs were combined for analysis. For this report, the team identified three themes in the dataset and clustered the coded data into categories and subcategories for each theme. Finally, they produced a rank-ordered list of categories and subcategories to identify highly salient (mostly frequently appearing categories/ subcategories) and relevant (mentioned in at least half of the FGs) by theme. This is based on the assumptions that frequently discussed issues are important to people, [21] and that recommendations for programs or policies should focus on the most relevant issues.

### Ethical considerations

The study was approved by the Medical Research and Ethics Committees at Queen’s University Belfast (Pre FREC Ref 19.24v3) and the Cross River State Ministry of Health’s Health Research and Ethics Committee (CRS/MH/HREH/019/ Vol.V1/175). All participants gave oral consent upon joining the study and re-confirmed it in writing before being interviewed. All procedures were carried out in accordance with the Helsinki Declaration.

### Steps to ensure rigour

The researchers applied the following procedures to ensure high-quality findings: following a well-designed research protocol, pilot testing the interview guide, carefully training the data collectors, maintaining an audit trail, having a multidisciplinary team co-develop codes and categories during data analysis (researcher triangulation), including an experienced qualitative researcher on the team, supporting assertions with evidence (quotes) and using the Standards for Reporting Qualitative Research (SRQR) criteria to evaluate the research process and methods. [22]

### Presenting findings

To distinguish between researcher and participant views, quotes appear in italics and are labeled with a unique anonymous ID, described above. Where needed, the analysts lightly edited participants’ statements to increase readability while retaining their original meaning. Ellipses (…) show where words were removed and text in square brackets indicate where they have been added.

## Results

### Study participants

Forty-four parents were interviewed in ten focus groups, 28 (63%) of them female. Participants ranged in age from 23-61 years (mean 41.3 ± 7.8 years) and had from one to seven children (mean 3.6 ± 1.3). Most participants were civil servants (civil or public servants, health workers, social workers, teachers,) or businesspeople (e.g., trader, seamstress, lottery ticket seller). Other occupations represented by very few participants include farmers, housewives, clergy or the unemployed. The focus groups ranged in size from two to eight members (mean = 4.4), half of them ‘urban’ and the other half, ‘rural’. All but one group consisted of female and male members (Table 1).

**Table 1.**
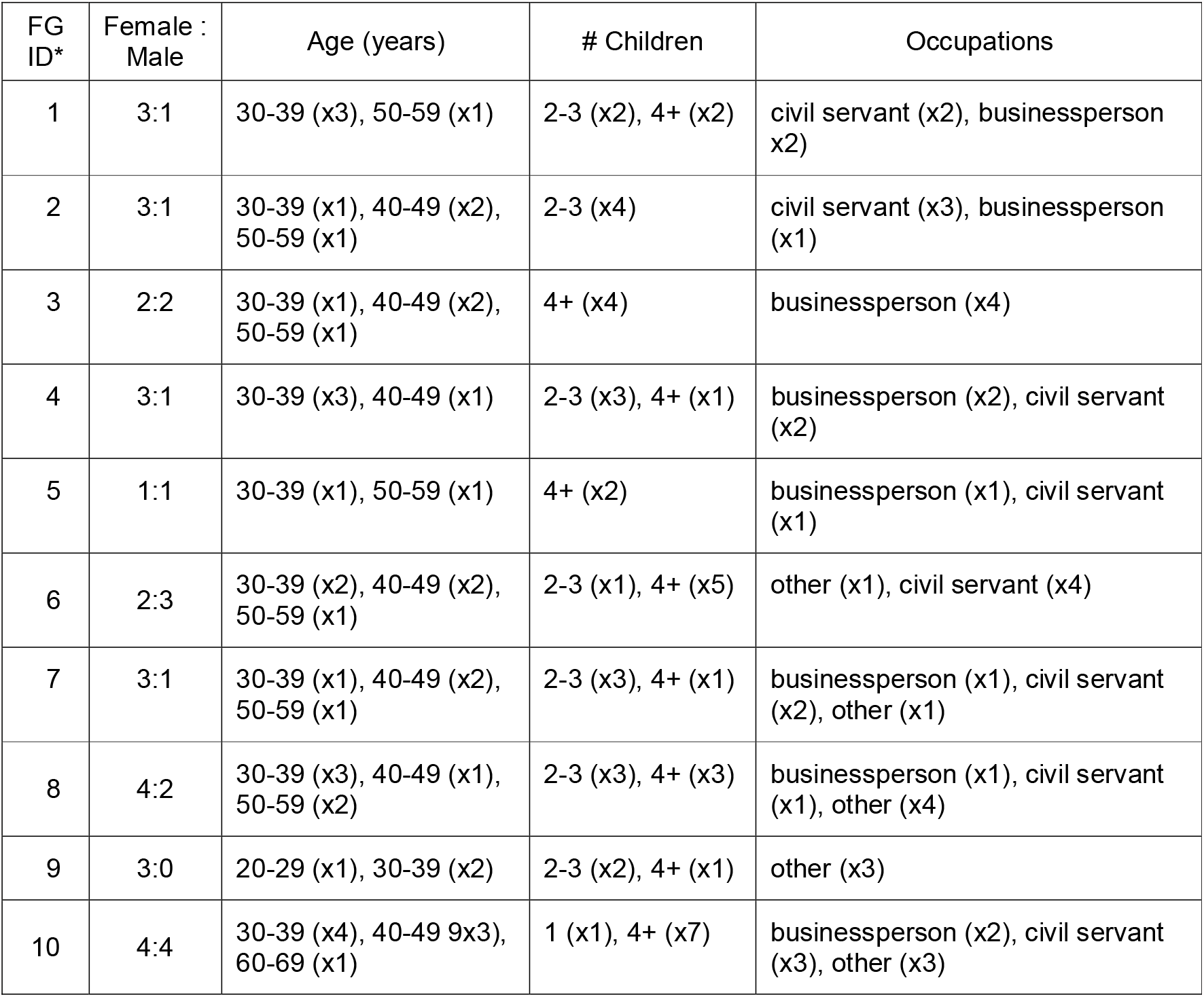
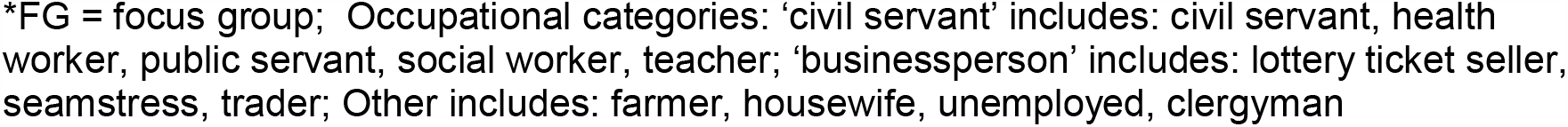
Characteristics of parents of children who failed a school vision screening test and did not bring the child for follow-up examination at a local hospital-based eye clinic (n=44 parents in ten focus groups)

| FG ID* | Female : Male | Age (years) | # Children | Occupations |
| --- | --- | --- | --- | --- |
| 1 | 3:1 | 30-39 (x3), 50-59 (x1) | 2-3 (x2), 4+ (x2) | civil servant (x2), businessperson (x2) |
| 2 | 3:1 | 30-39 (x1), 40-49 (x2), 50-59 (x1) | 2-3 (x4) | civil servant (x3), businessperson (x1) |
| 3 | 2:2 | 30-39 (x1), 40-49 (x2), 50-59 (x1) | 4+ (x4) | businessperson (x4) |
| 4 | 3:1 | 30-39 (x3), 40-49 (x1) | 2-3 (x3), 4+ (x1) | businessperson (x2), civil servant (x2) |
| 5 | 1:1 | 30-39 (x1), 50-59 (x1) | 4+ (x2) | businessperson (x1), civil servant (x1) |
| 6 | 2:3 | 30-39 (x2), 40-49 (x2), 50-59 (x1) | 2-3 (x1), 4+ (x5) | other (x1), civil servant (x4) |
| 7 | 3:1 | 30-39 (x1), 40-49 (x2), 50-59 (x1) | 2-3 (x3), 4+ (x1) | businessperson (x1), civil servant (x2), other (x1) |
| 8 | 4:2 | 30-39 (x3), 40-49 (x1), 50-59 (x2) | 2-3 (x3), 4+ (x3) | businessperson (x1), civil servant (x1), other (x4) |
| 9 | 3:0 | 20-29 (x1), 30-39 (x2) | 2-3 (x2), 4+ (x1) | other (x3) |
| 10 | 4:4 | 30-39 (x4), 40-49 (x3), 60-69 (x1) | 1 (x1), 4+ (x7) | businessperson (x2), civil servant (x3), other (x3) |

\*FG = focus group; Occupational categories: ‘civil servant’ includes: civil servant, health worker, public servant, social worker, teacher; ‘businessperson’ includes: lottery ticket seller, seamstress, trader; Other includes: farmer, housewife, unemployed, clergyman

### Themes

The three themes identified in the dataset are: modifiable barriers, contextual factors or obstacles a child eye care program cannot easily address, and participants’ recommendations on how to improve the eye care program. The most salient and/or relevant responses in each theme are listed in Table 2, with additional quotes covering the full range of categories and codes provided in Supplementary File 2.

**Table 2.** Highly salient and relevant barriers to parental adherence to referral for follow-up eye care for children failing a vision screening test in Cross River, Nigeria*

|  |
| --- |
| **Theme 1. Modifiable Factors**<br>Parental beliefs<br>*(about child’s vision/need for care)*<br>*(about costs at eye clinic)*<br>Problems with the referral letter<br>*(no letter given to parents)*<br>*(no explanation of letter contents)* |
| **Theme 2. Contextual Factors**<br>Parents’ situation<br>*(no money)*<br>*(cannot bring child to the clinic)*<br>Parental attitude towards children<br>*(uncaring)*<br>*(careless or forgetful)*<br>Parental beliefs about care<br>*(about hospitals)*<br>*(spiritual or religious beliefs)* |
| **Theme 3. Participant Recommendations**<br>Educate people<br>*(educate the public)*<br>*(correct misconceptions)* |

\* Findings in each theme are organized by category and sub-category (in parentheses). Within each theme, highly salient issues comprised at least 50% of all responses, and highly relevant ones were discussed in at least half of the focus groups.

### Theme 1: modifiable barriers

Parental beliefs, or views held as true or false without evidence or proof, [22] were a major barrier to referral adherence according to our study participants. Key among them was the belief that one’s child did not have a vision problem. As one mother explained, *“I felt there was nothing wrong with his eyes because he’s never complained to me* [so] *I didn’t take it serious”* (FG4-F1). Many parents also had the misperception there was a cost associated with follow-up care at the local eye clinic identified in the referral letter. One woman described this situation, saying *“I saw the paper my child brought home and thought it was something I needed to pay money for so I said we wouldn’t be able to go”* (FG3-F1). Another woman noted that *“many parents … think it will be expensive”* (FG9-F1) and so choose not to attend. Participants also explained that either their child had not brought the referral letter home (*“When you give them the referral letter* [many children] *won’t give it to their parents”;* FG8-F1) or had not explained the contents of the letter to them (*“If he* [referring to her son] *had informed me, I would have taken him there by now”*; FG7-F2).

### Theme 2: contextual factors

Although contextual factors lie outside the purview of a health care program, they can affect its acceptability, uptake, scalability and sustainability. Therefore, it is important to understand additional factors that can also be obstacles for parents seeking follow-up eye care for their children. Key among these factors were the overall family or parental situation, parents’ attitudes towards children or their beliefs about care.

Participants in several focus groups identified poverty as the main barrier to seeking follow-on eye care for their children (*“At the time there wasn’t even food for us to eat … that’s why we didn’t go* [to the eye clinic]*”*; FG7-F2). Other parents mentioned logistical difficulties, such as the lack of time (*“Because I didn’t have the time, I turned a deaf ear to it”;* FG2-F2) or an inability to take time off work (*“It all boils down to time. Some parents have very tight schedules –* [like me]. *At times I can be in the office from morning till midnight!”;* FG6-M3).

Examples of barriers due to parental attitudes, or their general predisposition to something as negative or positive, [22] include their being *‘careless’, ‘negligent’, ‘forgetful*’ or *‘disinterested in their children’*. As one woman explained: “[Some parents] *don’t value their children because they have so many”* (FG5-F1). In other cases, children are cared for by a person other than the parents *“who maybe isn’t really interested in the child’s health”* (FG5-F3).

In other cases, parents are *“scared to take their children to the hospital* [because] *they’re afraid their children’s eyes will get worse”* (FG10-M3). Still others believe *“going to the hospital is taboo”* or that the hospital is only *“for big people* [or] *people that inherit sickness”* (FG2-F3). Many Pentecostal or charismatic churches do not endorse the use of allopathic medicine and instead exhort members to only rely on the power of prayer and/or laying on of hands to cure health problems. As one man explained, *“Some of them* [parents] *don’t believe in medical treatment*, [because of] *their orthodox church”* (FG1-M1).

### Theme 3: participant recommendations

One of the main recommendations was to educate the public about children’s eye health. As one group participant noted, *“I think it* [not seeking follow-on care] *may be due to the lack of information … a parent may not be informed on how serious the issue of that child is”* (FG4-F1). In a different group, another parent thought that *“some parents don’t have knowledge about all this”* (FG8-M1). Another recommendation was to specifically address misperceptions held by parents, such as thinking they would have to pay for the appointment or that the healthcare team would return to the school to provide follow-up care at a later date. As one father explained, *“I didn’t go* [to the clinic] *because* [I thought] *the team will come back to the school for further care”* (FG10-M4). Some parents also noted it is important to ensure the referral letter looks official because *“some people believe only government officials are supposed to go into schools for immunisation or eye treatments – anything to do with children’s health”* (FG4-M1).

## DISCUSSION

Our study aimed to learn why some parents do not adhere to referrals for follow-up eye examinations for children failing a vision screening test in Nigeria. The two main modifiable barriers were parents’ beliefs about their children’s vision and anticipated costs at an eye clinic, and problems receiving information about the referral for follow-up care. Contextual factors that could reduce parents’ acceptance of the child vision care program in Cross River State include parents’ situation (poverty and the inability to bring the child to the clinic), overall negative attitudes towards children, and beliefs about hospitals and non-allopathic care. Recommendations to improve the program centred around educating the general public and correcting parents’ misperceptions.

As in other studies, the present research identified several factors affecting adherence to referral for follow-up eye care for children who fail a school vision screening test. A Brazilian study found a significant number of parents did not bring children for follow-up examinations even when free transportation, free spectacles and appointments on weekends were offered. [24] Reasons for not attending the follow up appointment included not knowing about it or forgetting an appointment, as well as not having someone to look after the child’s siblings or thinking the child did not have a vision problem.

Other studies found that parents generally only seek help only for noticeable ocular symptoms such as watering or discharge from the eyes, reddened or itchy eyes, [25,26] or for health problems they believe are severe. [27-29] This is also the case for adults, whose search for eye care is generally symptom-driven. [30,31] Given that children with RE do not exhibit such signs, parents in Nigeria and other countries do not seek additional eye care for their children, even when provided a referral letter recommending that they do so. [32-34] In some communities, there is also a common belief that eye problems only occur in adults, a view that was expressed in the current study and elsewhere.[24] Perhaps of more concern is the fact that several studies show that eye health is not a priority issue for the public. [35] These findings point to the more general issue of low health literacy or awareness about ocular health issues such as VI or URE. [36] This may help explain why parents do not understand the need for a follow-up eye examination for their children or additional care after failing a vision screening test.[37]

Other parental barriers include a lack of cooperation or interference from family members who hold differing beliefs about when and where to seek care for a child. In Africa, decisions about when and where to seek care outside the household is usually made by a “therapy management group” [38] consisting of key persons in the immediate family or household plus others who are part of the extended family or seen as particular wise or influential. This explains why mothers may want to bring a child for follow-up vision care yet defer to opposing views held by the child’s father or other decision makers. [39,40] This points to the need to identify and target key decision-makers in a child’s therapy management group when offering household- or community-based health education campaigns.

In virtually all communities, there are multiple options when seeking care for a health problem, from home-based advice and folk remedies, secular and spiritual healers, and biomedical practitioners and services – what is referred to as ‘medical pluralism’. In such settings, misconceptions about the causes of eye problems and VI plays an important role [41,42] may lead people to use traditional remedies or buy medicine from a pharmacy or drug peddler without a formal diagnosis.[43] In some cases, people may assume that vision problems are caused by witchcraft or are members of a strict orthodox (Pentecostal) church and so reject allopathic treatment. There are also beliefs about hospitals that can prevent people from seeking care in hospital-based eye clinics as reported in the present study.

These barriers are strengthened when eye care programs do not effectively communicate with parents to increase their understanding of URE and other invisible eye problems and the need for screening plus follow-up vision exams. In the present study, schoolchildren are expected to bring home a referral letter describing the need for a follow-up examination and where the local eye clinic is located. They are also supposed to explain the letter contents in such a way as to overcome barriers to action such as parental beliefs about vision and the importance of good eye care. Thus, although other studies have found an increased rate of follow-up exams when written notice of the follow-up referral is sent to parents, [44,45] how this is done may need to be tailored to fit local beliefs and practices.

As noted earlier, URE, the most common cause of VI in children, is most prevalent in low-income countries and communities. It is not surprising, therefore, that in this study and also prior research economic and/or logistical reasons can be major barriers to attending follow-up eye care appointments. [46,47] In the current study participants indicated that many parents in the area mistakenly believed that follow-up care in a local hospital would be expensive, which deterred many of them from adhering to the referral for such services.

Logistical barriers identified in this and earlier studies include organizing appointment times around busy work schedules, taking time off work leading to lost wages, [43,48] not having an adult available to accompany the child to a clinic [24] plus finding and paying for transport. [49,50] In some cases, these problems are amplified by facility-related issues such as long waits at clinics, inconvenient operating hours, and negative attitudes of practitioners towards their patients, [24,43] all of which were mentioned by participants in the present study.

Several authors of prior studies about ocular and other health issues recommend offering health promotion campaigns to reduce misconceptions among parents and the general public, [30,31] addressing such issues as the difference between vision screening and comprehensive eye tests. [51-53] Such information campaigns must also address long-standing beliefs and underlying cultural and religious factors. [54]

Some researchers have suggested simplifying referral pathways between the education and health systems, such as by involving teachers or school nurses in screening and/or communicating with parents. [55,56] Another recommendation is to personalize communications with parents, such as directly calling parents before the first follow-up appointment [57,58] and providing condition-specific information about their children’s condition. It is also important to identify locally popular communication channels through a participatory process with local community members, especially parents of young children. [59] Messages should be targeted to specific barriers that are prevalent in the target communities and co-developed with school-aged parents. [60]

Logistical support can also help more parents bring children for follow-up eye exams. Examples include providing parents with reminders and help scheduling appointments [30] or providing follow-up care in schoolyards with mobile eye care units. [31] Such suggestions would need to be pilot tested and modified to fit local conditions and expectations.

### Strengths and weaknesses

A strength of this study is having used a qualitative approach allowed the researchers to provide insight into the beliefs, attitudes and practices of parents that they themselves linked to not adhering to a referral for follow-up eye care for their children. There is growing consensus on providing care that is based on the views of patients and their families.

Qualitative research methods are ideal for understanding their experiences. [61] Health care programs, including those to protect children’s vision, are more likely to succeed if they are based on local evidence regarding people’s attitudes, beliefs and behaviours. This includes identifying ways to promote the uptake of services. [62] However, there is considerably more funded research using qualitative and/or mixed methods approaches than what is being published in ophthalmology and vision science journals. [63] The present study, which was rigorously designed and conducted, provides new understandings for the Nigerian government.

One weakness of this study is that it did not follow standard steps to determine sample size, which is usually determined by conducting preliminary analysis of the data while still in the research setting and able to continue collecting data. This is referred to as saturation or having enough data to answer the research questions. However, although saturation is considered a gold standard in qualitative research, there are various ways to achieve it. Saturation was developed for use in Grounded Theory studies and refers to “theoretical saturation”. The aim is to analyse data inductively (not based on pre-existing categories or themes) while still collecting data to ensure there is enough information to create theoretical categories that underpin a new model or theory. The aims and methods used in such studies are quite different from those of a descriptive qualitative study that included a mainly deductive analytic approach (i.e., sorting data into pre-existing topics drawn from the interview guide). Evidence for having reached “*a priori* thematic saturation” [64] exists in the analysts’ having reached consensus on themes and categories that are both highly salient (frequently mentioned) and relevant (identified in at least half of all groups) supported by the raw data.

A second weakness is not finding differences in the views of parents in urban and rural schools. This may be an artefact of not collecting enough data to identify differences between these two sets of parents. However, it may also be that the underlying beliefs, attitudes and practices regarding children’s vision and eye health are in response to a single set of underlying cultural, socioeconomic and health system factors that do not markedly differ in these two contexts. Further research could be done in other parts of Nigeria, where different ethnic/language groups live to see if lessons learned in Cross River State can be applied to other parts of Nigeria or Sub-Saharan Africa.

## Supporting information

Supplemental Interview Guide

Supplemental Table of Quotes by Category

## Data Availability

The data for this study consists of verbatim transcripts in English from 10 focus group interviews with 44 parents of schoolchildren who failed a vision screening test. None of the participants consented to have the transcripts made publicly available. Given that thd transcripts contain sensitive data that may potentially reveal the identity of focus group members, data availability is restricted for ethical reasons. On request, the corresponding author (Dr Lynne Lohfeld) can make condensed meaning units or parts of transcripts available after text has been de-identified.

## Acknowledgements

The authors would like to thank the parents in Cross River State, Nigeria, who participated in the focus group discussions, and the fieldwork team who collected and prepared the data for analysis: Mrs Okwoalice Ita with Drs Kenneth Azubike, Emmanuel Iwong and Emmanuel Odor.

## Conflict of Interest

Ving Fai Chan is a Trustee of Vision Aid Overseas, a non-governmental organization delivering refractive services in LMICs, including Africa. Nathan Congdon is the Director of Research for Orbis International, which delivers eye care, including children’s refractive services, in Africa and other low-resource settings. No other author had no conflict of interest to declare.

## Notes

### Funding Statement

This project was funded by a Northern Ireland Department for the Economy (NI DfE) Global Challenges Research Grant (GCRF) awarded to Dr Ving Chan. None of the authors or their institutions received payment or services from a thrid party for any aspects of the submitted work.

### Author Declarations

The study was approved by the Medical Research and Ethics Committees at Queens University Belfast (Pre FREC Ref 19.24v3) and the Cross River State Ministry of Health, Health Research and Ethics Committee (CRS/MH/HREH/019/ Vol.V1/175).

