## Supplemental Interview Guide for "*“You’re too young to have an eye problem”*: Reasons for nonadherence to referrals for follow-up eye care for schoolchildren in Nigeria -- a descriptive qualitative study"

Supplementary File 1.

Questions asked during focus group discussions with parents or guardians of children who failed the school-based vision screening test and did not bring the child for a recommended follow-up eye exam

1. Understanding of Vision Screening Test Results

1a. How many of you know that your child’s school offers free vision screening or an eye test for your children? [show of hands, to be counted)

1b. What were you told about the results of that eye test?

PROBE: Did the teacher tell you or did you receive a note from your child about the test results?

1c. What do you think that might mean for your child’s health and wellbeing now?

1d. What about in the future?

2. Understanding the Value of Further Testing Children Who Fail a Vision Screening Test

2a. After your child had his/her vision tested at school, what were you told you should do about your child’s eye health?

PROBE: Why do you think further testing was recommended for your child’s eyes?

2b. We know that some parents or guardians of children who are referred for further examination at the eye clinic choose not to do so. What do you think are the main reasons some families make that choice?

3. IDENTIFY TYPICAL OR PREFERRED PRACTITIONER

3a. Why do you think some families choose to bring their children in for an eye examination?

PROBE FOR FULL SET OF REASONS BY ASKING: Anything else? Or, any other reasons that you can think of? REPEAT THE LIST OF REASONS FROM YOUR NOTES AND WAIT FOR FURTHER RESPONSES.

PROBE: Who in the household generally makes such decisions?

4. WRAP UP FINAL THOUGHTS & DEMOGRAPHIC/BACKGROUND INFORMATION

4a. We’re almost at the end of our meeting today. Thinking back to everything we talked about, is there anything you think I left out or didn’t ask you that would be important to know?

4b.	Is there anything you would like to talk about in more detail?

4c.	I want to bring back your key messages to the other members of our team, so I’ll ask each of you, one by one: What is your key message that you’d like me to bring to the others?

4d.	Is there anything else you’d like us to know before we end our meeting?

Collect demographic information from each participant.

Note: For sections 1-3, provide a brief summary of points raised and ask participants if they want to modify or add to their information. Include facilitator and note-taker debriefing to audio file of focus group discussion.
