## Supplemental Table of Quotes by Category for "*“You’re too young to have an eye problem”*: Reasons for nonadherence to referrals for follow-up eye care for schoolchildren in Nigeria -- a descriptive qualitative study"

Supplementary File 2. Additional quotes, by theme and category, from parents of children who failed a school-based vision test and did not adhere to the referral for additional care at a local hospital-based eye clinic

Theme 1: Modifiable Barriers

| CATEGORY: Parental Beliefs about … | | QUOTES |
| --- | --- | --- |
| on child’s vision &  need for care | there is no problem | FG9-F1: *“Another reason is disbelief* [denial]. *Some people don’t believe that anything* [is wrong with the child’s eyes]. *Like,* [with] *some children who lived nearby in my neighbourhood, when their eyes were tested their parents said, ‘God forbid, it’s not my child* [who has a problem]!’ *So, they just leave it.”*  ***IWer: “Is it not believing in the test, or that the child has a problem?”*** *“Not believing there’s a problem. They will say there’s no need for further testing because there’s no problem.”*  FG3-F2: *“Some parents might say that all this time my child has been walking around without any problem with his eyes; then why do they* [the school] *say she has a problem?”*  FG1-M1: *“Some ignorant parents may think that it is just ‘eehh’;* [indicating ‘not very important with a shrug]. *It’s ignorance on the part of the parents; they’re unaware of what’s going on.”* |
|  | eye problems not in children | FG8-F1: “[With a young child] y*ou will not even take anything serious you will relax. That’s the reason.”* |
|  | not bad enough to take action | FG1-F1: *“In my case, since my child wasn’t complaining about a physical problem, I felt it may not be important for me to go there. There wasn’t any eye drooping and he [referring to her child] was going about with his normal business so I felt it isn’t that serious, and I overlooked it.”* |
|  | spectacles | *“Glasses are for adults and not children.”* (FG10-F2)  *“I was afraid that if I started wearing glasses it can negatively affect my eyes.”* (FG10-M3)  *“I told my father* [about my vision problem] *and he said, ‘Okay, use my General Hospital card and go to the General Hospital for testing’. But I said, “Papa, leave it. They’ll just recommend eye glasses for me and I don’t want to wear glasses.”* (FG10-F4) |

Table continued on next page

| CATEGORY: Parental Beliefs | | QUOTES |
| --- | --- | --- |
| on need for care | other issues | *“Some parents would feel if they go* [for additional care] *it will worsen the situation of their children.”* (FG4-M1) |
| CATEGORY: Parental Beliefs about … | | QUOTES |
| about cost | thinking they’d have to pay | *“I was concerned about going* [as] they [would be]) *asking me to buy* – [they would tell me] *I must pay for the drugs, I have to pay for the glasses, and all that*.” (FG4-F1)  *“If you find this sort of thing* [it’s because even though] *the person says, ‘I should go’, but they may think it’ll cost money and so they don’t want to go there because of the possible financial costs.”* (FG5-F1) |
|  | thinking it was expensive | “*Sometimes some parents are scared that the cost for eye treatment is very high … and may think they* [hospital staff] *will demand a lot of money but they don’t have that kind of money to able to pay the hospital bill. I think that’s one of the major challenges.”* (FG10-M1)  *“I’ve been postponing* [taking my son for an eye exam] *thinking that the cost might be high.”* (FG5-F1) |
|  | always have to pay something at hospital | “*As far as Nigeria is concerned … less privileged people cannot go to hospital. Even if a free test is done [like at the school] then they’re asked to go to the hospital. But they know that the nurses and the doctor won’t pay attention to them if they don’t bring money. First, they ask you to buy* [a patient registration] *card. Then they will prescribe drugs for you and ask you to buy* [them]. *They will not even start treating you without you paying money!”* (FG2-M1)  *“Most parents will think if they go there, the hospital will charge them.”* (FG9-F3) |

Table continued on next page

| CATEGORY: Issues with referral system | | QUOTES |
| --- | --- | --- |
| problems with referral letter | no referral letter given to parents | *“My children came – especially my last son – and told me that they’d tested him. What made me not go [to the hospital] with the child is that I didn’t receive any note from them* [the school].” (FG10-F3)  “Because my son didn’t tell me [he failed the school vision test] and I didn’t see the [referral] letter or I would have gone to the hospital for screening. I have gone to the hospital they have screened me.” (FG7-F2) |
|  | contents not explained to parent | *“In my case, my son didn’t tell me* [about the referral to hospital]. *Had he told me, I would have gone – I would have taken him to the hospital* [for the follow-up eye exam].” (FG7-F3)  “Some parents won’t understand what they were asked to do.” (FG5-M1) |
| problem with referral letter | letter does not look official | “[I didn’t go with my child to the hospital] *because I didn’t get* confirmation from the authority, from the school authority to go for further test, that’s why I couldn’t go for the eye test.” (FG10-M4)  “[My children said] *we’re to go to the general hospital for further checks and to get some glasses, which I didn’t agree to because I didn’t hear that from the authorities.”* (FG10-M3)  *“If something isn’t from the government … if it’s from a non-governmental organization and nothing is attached to it to let people know that it’s something important* [and endorsed by the government] *… people will be afraid. Some people believe only government officials are supposed to go into schools for immunization or eye treatments, or anything to do with children’s health.”* (FG4-M1) |

Table continued on next page

Theme 2: Contextual Factors

| CATEGORY: Financial Issues | | QUOTES |
| --- | --- | --- |
| no money,  poverty | general poverty | *“Poverty is, I mean, an understatement.”* (FG10-F1)  *“Simply put, a lot of our people are poor, and poverty affects our reasoning; you run around looking for things to eat. And it’s only later that they think about other problems that will bring them sorrow later… poverty is a problem* [implying the underlying problem].” (FG5-M1) |
|  | no money for hospital | *“Then, my problem was that I was not having any money by then to take her to the general hospital there, if not I could have will to be there.”* (FG9-M2)  *“For some parents it’s their financial status that will make them not to go further – not go to the hospital.”* (FG10-F3) |
|  | no money for transport | *“The reason why I didn’t go with her is because there was no transport.”* (FG7-F2)  “[Another reason people do not go for follow-up care] *is the lack of funds. When they* [the family] *check their financial status, they say, “I don’t even have money for transportation, how will I go to the hospital?”* (FG10-M1) |
|  | no money for the care | *“Some will think about money. Because the most important thing is money. There is no way you will go and you won’t be spending money. But, if the money isn’t there you will feel very reluctant to go. And in some cases after the test, they will tell you, “Go and do this test.” And you have to pay money!* [She threw her hands open and opened her eyes wide]. *So that discourages some parents to continue* [testing their child’s vision].” (FG2-F1)  *“Well, in a situation like this, I don’t see any reason that somebody* [parent or guardian] *would notice their child is sick and wouldn’t get treatment for them. But … if somebody doesn’t have the money, he might decide to do nothing about it because he has no money.”* (FG3-M2) |

Table continued on next page

| CATEGORY: Logistical Issues | | | | QUOTES |
| --- | --- | --- | --- | --- |
| making an Appointment | | no time | | *“Money is not the only reason. Maybe some parents are too busy.”* (FG9-F1)  *“After the* [screening] *test we were asked to go to the clinic, but I didn’t have time to go.”* (FG5-M1) |
|  |  | can’t take time off work | | *“My work didn’t permit me to….it didn’t give me free time to take him there.”* (FG4-F1)  *“He* [referring to his son] *kept pestering me* [to go]. *I told him I don’t have the time because I leave home in the morning and can’t leave the office until evening. So, I told him we would go, but I didn’t* [take him].” (FG6-M3) |
|  |  | Scheduling Issue | | *“We know that when the time comes to do an eye check-up for children, some people won’t go. They’ll have an appointment scheduled but might not be available* [at that time].*”* (FG10-F4) |
| CATEGORY: Illiteracy | | | QUOTES | |
| lack of education / knowledge | | | *“We still have a lot of illiteracy in town.”* (FG4-M1)  *“They* [referring to his children] *bring home letter for their mother, who can’t read. And she says, ‘Give me the letter and when your father comes* [home] *I’ll give it to him to read.’ By the time the father comes home, she’s forgotten so she doesn’t give the letter to him!”* (FG7-M1) | |
| CATEGORY: Parental Attitudes | | | QUOTES | |
| eye care not a priority | careless about vision care | | *“If a parent refuses to take their child for further care in this kind of situation, it’s probably due to carelessness.”* (FG5-F1) | |
|  | neglect | | *“For some it’s due to negligence.”* (FG4-M1) | |
|  | forgetting | | *“It escaped my memory -- even when I was thinking about it today,* [I was wondering], *will I be available to go* [to the clinic]? *And before the day* [of the appointment], *it escaped my memory.”* (FG6-F2) | |

Table continued on next page

| CATEGORY: parental attitudes | | QUOTES |
| --- | --- | --- |
| eye care not a priority | father’s attitude | *“My eldest son has the same eye problem that my husband does so … he feels like it’s nothing* [and] *when he’s [good and] ready he’ll take our son for treatment.”* (FG4-F2)  *“In a family if your husband says ‘No’* … *the child won’t go to the hospital.”* (FG9-F1) |
| children not a priority | not taking care of children | *“Some parents only give birth to their children, but don’t take care of them the way they’re supposed to.”* (FG8-M2)  *“Ehh, some children suffer a lot. Their parents are only interested in their children helping them, not the parents helping their children. If you were to spy into most homes, you’d want to become Jesus Christ so you could save the children from what they are going through. They don’t even eat well!”* (FG5-M1) |
|  | disinterested in children | *“Some of them just take things for granted. Most of them actually have the money to take the child to the hospital, but they just take everything for granted and just leave it alone.”* (FG9-F1)  *“Some parents may not be interested.”* (FG1-F2) |
|  | no empathy for others’ suffering | *“If you don’t love your children, as someone else said, you won’t even know if they’re in pain and you’ll wait until you see their health decline.”* (FG8-M2)  *“I can see that some people will be the end of their families. They don’t feel pain from things that don’t happen to them.”* (FG2-M1) |
|  | not my child | *“Some guardian or parents don’t take their children there* [referring to hospital] *-- mostly guardians* [of] … *their brother’s children. But because they’re not the person who gave birth to those children. So … they won’t to spend their money on the child because it’s not their biological children. For that reason they prefer to leave such children like that and allow their eyes to get bad -- which is very bad.”* (FG8-M2) |

Table continued on next page

| CATEGORY: Parental Beliefs About Care | | QUOTES |
| --- | --- | --- |
| beliefs about hospitals | fear | *“I think one of the problems is that some parents may be scared of taking their children to the hospital.”* (FG10-M3) |
|  | will cause more damage | *“Some* [parents] *could be afraid of further damaging their children’s sight.”* (FG10-M3) |
|  | receive a diagnosis & person becomes ill | *“A main reason why some people don’t go to the hospital is they don’t think they’re sick. But once they go to the hospital and they [the hospital staff] tell them they have a disease, then they become sick… they believe that if someone mentions a sickness of some kind, they will get it so that makes some people afraid to go to the hospital.”* (FG5-M1) |
| need for prayer | pray first | *“Some parents are like me. When something happens, I first will handle the spiritual aspects before getting medical attention because I know there are some cases that were resolved that like in the past…* [gave example of a child in her parish that had an eye problem] *I prayed for the child and laid my hands [on him]. And when they took him for the* [eye] *test … his eyesight was restored and everything was normal. That’s why some of us believe that the spirit controls the physical. It’s not that we don’t believe in the power of modern medicine, but sometimes it* [the problem] *can be a spiritual attack.”* (FG10-M4) |
| traditional remedies | use traditional remedies | *“I was taking care of a blind man and asked him, ‘How did you become blind?’ He said, ‘It’s because of measles.’ He had measles when he was young and his parents put drops of a traditional remedy in his eyes. ‘Why? I asked him. He said, ‘They didn’t know they should’ve taken me to the hospital’.”* (FG10-F1)  *“Some people believe in doing that* [giving traditional remedies] *at even a tender age.”* (FG10-F2) |
| religious beliefs | Apostolic churches forbid allopathy | *“It may be because of their religion. Like, the Apostolic faith; they tell you, ’Don’t use Western medicine’.”* (FG2-F1) |
|  | divine intervention | FG10_M3: *“Since I was one year old until I was 10 … I couldn’t see anything. In my 10^th^ year … I had a dream where I was taken to the hospital and, operated on. Then I woke up and started seeing when I was 10. So, since then I haven’t gone to any medical* [person] *or received any major medical treatment.”* (FG10-M3) |
| spiritual beliefs | | *“Sometimes it’s not a financial issue but a problem from attack* [witchcraft] *and then* *we need to fasten up in prayers* [seriously pray]” (FG10-F4). |

Table continued on the next page

| CATEGORY: Parental Beliefs About Care | | QUOTES |
| --- | --- | --- |
| seek care only when very sick | | *“It’s very rare to see adults around here going on their own* [initiative] *to the hospital for a check-up unless they’ve been diagnosed with BP, a heart problem or diabetes – a chronic condition. It’s just not common around here. For instance, until last month it was over a year since I’d had my BP checked!*” (FG6-M1) |
| CATEGORY: Facility-related Issues | | QUOTES |
| clinic or hospital issue | long wait | *“You go there* [to the hospital] *but don’t get to see the doctor… That’s stressful for parents because they have things to do at home rather than going to the hospital and wasting the whole day* [waiting to see the doctor].” (FG2-F1) |
|  | staff attitudes | *“Doctors have to show sympathy. Some doctors, they don’t have that human sympathy.”* (FG2-F1) |

Theme 3: Recommendations for Strengthening Children’s Eye Care Program (made by participants)

| CATEGORY: Focus on people | | QUOTES |
| --- | --- | --- |
| increase health literacy | educate the public | *“The biggest barrier is ignorance.* [People say], *‘If they really intend to treat my child, they should give us a date and come back to the school* [and provide treatment there].’ *I think that’s seriously ignorant! My opinion is ignorance is the main reason.”* (FG10-M1)  *“They* [referring to underprivileged or poor families] *may not have a radio or a television … to listen to or watch, where they would hear information like this.”* (FG1-F2) |
|  | correct misinformation for parents | *“My children brought the referral letters to me, but I didn’t follow their advice. I followed up with* [an appointment at the] *General Hospital, where my doctor promised me that the people* [offering eye care] *will come back by September. So, I let the children wait.”* (FG10-F1) |
